# Machine learning and biological validation identify sphingolipids as potential mediators of paclitaxel-induced neuropathy in cancer patients

**DOI:** 10.1101/2023.10.08.23296716

**Authors:** Jörn Lötsch, Khayal Gasimli, Sebastian Malkusch, Lisa Hahnefeld, Carlo Angioni, Yannick Schreiber, Sandra Trautmann, Saskia Wedel, Dominique Thomas, Nerea Ferreiros Bouzas, Christian Brandts, Benjamin Schnappauf, Christine Solbach, Gerd Geisslinger, Marco Sisignano

## Abstract

**Background:** Chemotherapy-induced peripheral neuropathy (CIPN) is a serious therapy-limiting side effect of commonly used anticancer drugs. Previous studies suggest that lipids may play a role in CIPN. Therefore, the present study aimed to identify the particular types of lipids that are regulated as a consequence of paclitaxel administration and may be associated with the occurrence of post-therapeutic neuropathy.

**Methods:** High resolution mass spectrometry lipidomics was applied to quantify d = 255 different lipid mediators in the blood of n = 31 patients drawn before and after paclitaxel therapy for breast cancer treatment. A variety of supervised statistical and machine-learning methods was applied to identify lipids that were regulated during paclitaxel therapy or differed among patients with and without post-therapeutic neuropathy.

**Results:** Twenty-seven lipids were identified that carried relevant information to train machine learning algorithms to identify, in new cases, whether a blood sample was drawn before or after paclitaxel therapy with a median balanced accuracy of up to 90%. One of the top hits, sphinganine-1-phosphate (SA1P), was found to induce calcium transients in sensory neurons via the transient receptor potential vanilloid 1 (TRPV1) channel and sphingosine-1-phosphate receptors.SA1P also showed different blood concentrations between patients with and without neuropathy.

**Conclusions:** Present findings suggest a role for sphinganine-1-phosphate in paclitaxel-induced biological changes associated with neuropathic side effects. The identified SA1P, through its receptors, may provide a potential drug target for co-therapy with paclitaxel to reduce one of its major and therapy-limiting side effects.

## Introduction

Paclitaxel is a standard adjuvant treatment for breast cancer and several other cancers. Originally isolated from the yew tree *Taxus brevifolia*, it inhibits mitosis by stabilizing microtubules and preventing tubulin depolymerization (1–3). A serious dose- and therapy-limiting side effect, which it shares with other commonly used cytostatic drugs, is the chemotherapy-induced peripheral neuropathy and neuropathic pain (CIPN), which affects up to 80% of treated patients (4, 5). Patients report a variety of primarily sensory symptoms, encompassing sensations like numbness, paresthesia, spontaneous pain, and heightened sensitivity to mechanical and/or cold stimuli in their hands and feet. In more severe instances, the loss of vibration sense and joint position sense can further impact their functionality (6). Engaging in essential daily activities becomes challenging for patients, leading to difficulties in tasks like fine finger movements (such as buttoning clothing). Walking can induce pain due to mechanical hypersensitivity, while handling tasks like retrieving items from a fridge or exposure to cold weather may exacerbate symptoms (cold hypersensitivity). Chemotherapy can also emerge post-treatment in a state termed “coasting” where mild neuropathy can worsen, or new instances of CIPN may develop (7). Currently, there are no pharmacologic treatments for CIPN expert for duloxetine (4, 8, 9). Therefore, research on the mechanism of paclitaxel induced CIPN with possible identification of novel treatments is an active research topic.

Several genes and neurofilament proteins have been implicated in paclitaxel-induced neuropathy (10–12). More recently, lipid mediators have been shown to be produced at high levels in sensory neurons, neuronal tissue and immune cells after chemotherapy due to oxidative stress and have been shown to contribute to chemotherapy-induced neuropathy and neuropathic pain by modulating neuronal ion channels (13–16). Therefore, they are of particular interest as signaling molecules and potential markers for chemotherapy-induced neuropathy in patients. In fact, lipids are already considered markers for other neurological diseases such as Alzheimer’s disease and amyotrophic lateral sclerosis (ALS) (17–20). However, a systematic approach to investigate lipids in patients with paclitaxel-induced neuropathy has not been performed.

In this prospective clinical cohort study, plasma concentrations of 255 different lipid mediators were evaluated for changes in their concentrations associated with paclitaxel treatment. A comprehensive LC-MS/MS-based targeted and LC-QTOFMS-based untargeted lipidomics screening was performed on plasma samples from paclitaxel-treated patients. Lipid groups measured included eicosanoids, endocannabinoids, oxidized linoleic acid metabolites, sphingolipids, lysophospholipids and free fatty acids, many of which have been previously associated with persistent pain states (21–23). A data-driven approach was used to identify lipid mediators whose concentrations could be used to train machine learning algorithms to identify, in new cases, whether a plasma sample was collected before or after therapy, or from a patient with or without post-therapy neuropathy. The biological relevance of the findings was then validated in vitro by applying SA1P to primary sensory neurons using calcium imaging.

## Materials and Methods

### Patients and study design

This was a prospective single-arm study enrolling patients with breast cancer. The study was conducted in accordance with the Declaration of Helsinki on Biomedical Research Involving Human Subjects and was approved by the Ethics Committee of the Medical Faculty of the Goethe-University, Frankfurt am Main, Germany. Informed written consent was obtained from each of the participants.

Sixty patients (one male, 59 females with breast cancer and undergoing paclitaxel treatment were recruited from the Tumor Center of the University Hospital Frankfurt, Germany (UCT). Most patients (n = 47) received the “paclitaxel-weekly” schedule, consisting of 12 cycles of paclitaxel treatment (80 mg/m², each week); a few patients (n = 13) received mixed carboplatin/paclitaxel treatment (Table S1). A blood sample was collected from each patient before and after chemotherapy, and the degree of neuropathy after chemotherapy was assessed as described below. Plasma was isolated from the blood samples immediately after blood collection to ensure lipid stability. Plasma was stored at -80°C until analysis.

All patients provided a blood sample before chemotherapy; however, only 36 patients of the patients provided a second blood sample after chemotherapy. For our analysis, we focused on patients that had two blood samples. Therefore, 72 samples from 36 patients (before and after chemotherapy) were analyzed using both LC-MS/MS-based targeted and LC-QTOFMS-based untargeted lipidomics. A total of 255 individual lipids were detected in each sample. From the resulting data, five patients had to be excluded due to incomplete lipidomics data (more than 20% of the analytes could not be detected). The remaining 62 samples from 31 patients were used for the machine learning analysis (Figure 1).

**Figure 1:**
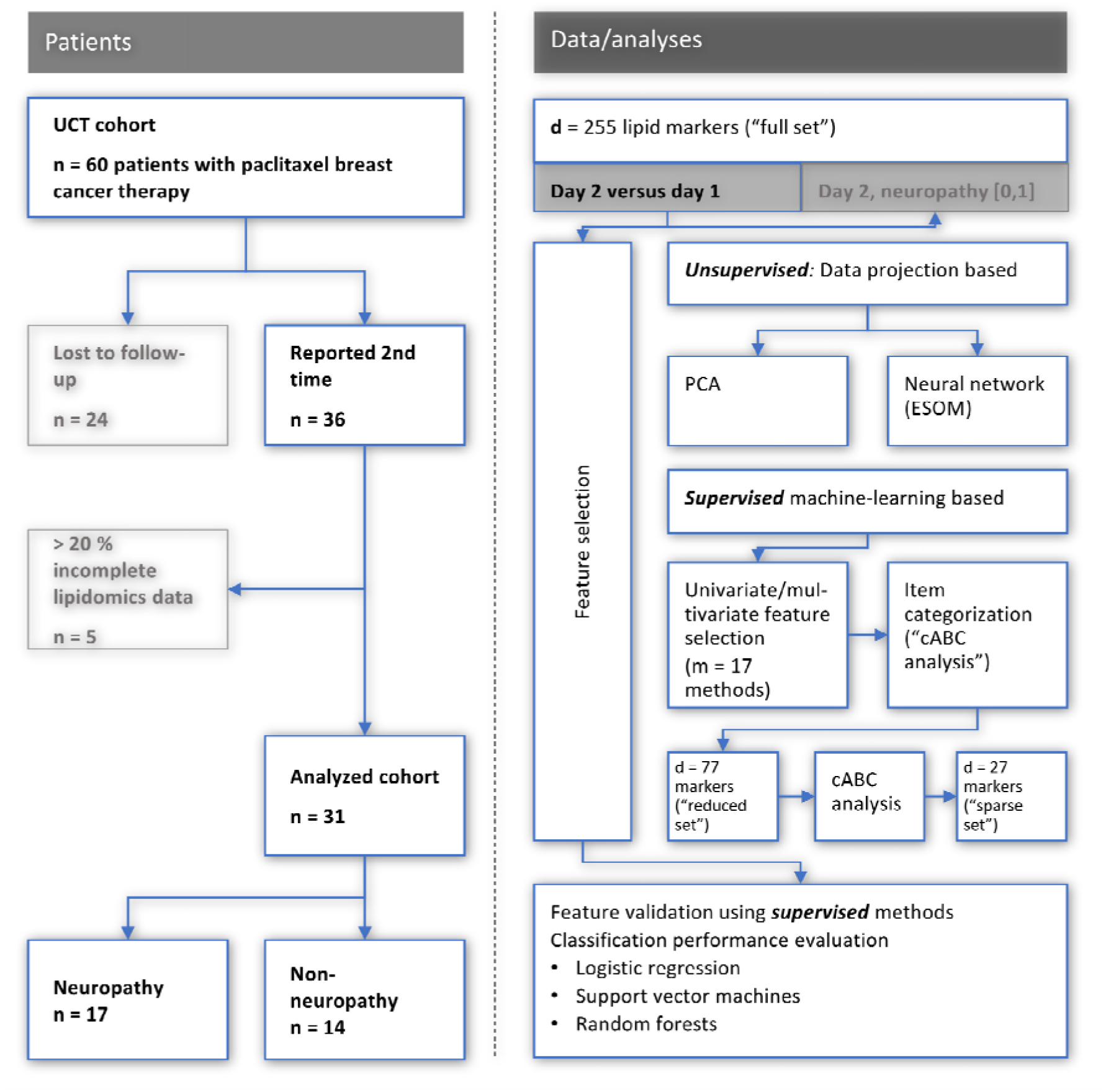
Flowchart showing the number of patients included and the workflow of the data analysis. UCT: University Cancer Center Frankfurt, PCA: principal component analysis, ESO: emergent self-organizing maps, cABC analysis: computed ABC analysis. The figure was created using Microsoft PowerPoint® (Redmond, WA, USA) on Microsoft Windows 11 running in a virtual machine powered by Virtual Box 6.1.36 (Oracle Corporation, Austin, TX, USA) as guest on Linux, and then further modified with the free vector graphics editor “Inkscape (version 1.2 for Linux, https://inkscape.org/

### Assessment of neuropathy

The occurrence and severity of peripheral neuropathy was assessed according to the guidelines of the NCI Common Terminology Criteria for Adverse Events (CTCAE) v5.0, Published: November 27, 2017, by the U.S. Department of Health and Human Services (https://ctep.cancer.gov/protocoldevelopment/electronic_applications/docs/ctcae_v5_quick_reference_5x7.pdf). Neuropathy assessment was performed prospectively, i.e., before the first paclitaxel treatment and again after the 12^th^ treatment cycle. Neuropathy was assessed regularly upon visit of the patient. The last assessment was performed 4.5 years after initial chemotherapy (Table S1).

The severity of neuropathy was graded into five grades: Grade 1 Mild; asymptomatic or mild symptoms; clinical or diagnostic observations only; intervention not indicated. Grade 2 Moderate; minimal, local or noninvasive intervention indicated; limiting age-appropriate instrumental ADL (activities of daily living). Grade 3 Severe or medically significant but not immediately life-threatening hospitalization or prolongation of hospitalization indicated; disabling; limiting self-care ADL. Grade 4 Life-threatening consequences; urgent intervention indicated. Grade 5 Death related to AE. In the present cohort, grades 1-3 were detected following paclitaxel chemotherapy (Table S1). Of the 31 patients with a full set of samples, 17 had neuropathy after chemotherapy (54.9%), 12 had grade 1, 3 had grade 2 and two patients with grade 3 (Table S1).

### Lipidomics analysis using LC-MS/MS and LC-QTOFMS

Blood was collected from patients in EDTA tubes and immediately centrifuged at 2000 xg for 10 minutes at 4°C.The supernatant was immediately frozen at -80°C until further processing. Approximately 2-3 ml of plasma was considered sufficient for each patient. Liquid chromatography-tandem mass spectrometry (LC-MS/MS) analysis of eicosanoids, oxidized linoleic acid metabolites (O(x)LAMs), prostanoids, endocannabinoids, LPAs, pterins, sphingolipids and ceramides, and lipidomics screening were performed as described previously (15, 22, 24). A total of 255 lipids were quantified in each plasma sample. These lipids belong to the groups of eicosanoids, oxidized linoleic acid metabolites, endocannabinoids, lysophosphatidic acids, pterins, sphingolipids, ceramides, cholesterols, cholesterol esters, diacylglycerols, triacylglycerols, phospholipids, lysophospholipids, and free fatty acids. Full details of the lipids detected are given in Table S2. Full details of the LC-MS methods used can be found in the Supporting Information Methods section (Tables 1-10).

### Data analysis

Programming was performed in R language (25) using the R software package (26), version 4.1.2, for Linux, available free of charge from the Comprehensive R Archive Network (CRAN) at https://CRAN.R-project.org/, and in Python language (27) using Python version 3.8.12, available free of charge at https://www.python.org (accessed March 1, 2022). The presentation emphasized as far as possible the presentation of raw data as advised in (28). The data analysis is summarized in Figure 1 and described in detail in the supporting information.

The data analysis combined statistical and machine learning methods in the sense of a so-called “mixture of experts” approach that has been shown repeatedly to be superior to relying on a single method (29, 30), such as regression analysis alone (31). The reason for the preference for using several but one method is that all statistical models inherently rely on underlying assumptions about a data set, some of which can be tested, but in practical scenarios it is often difficult to determine the best model to describe a real data set. Therefore, the results of the present analysis were derived from the consensus of several methods in an effort to increase their certainty. Of note, all statistical and machine-learning tests were performed two-sided, i.e. without a directed hypothesis, which was not available for all of the d = 255 lipid markers with respect to their regulation as an effect of paclitaxel and/or as a sign of the development of neuropathy. The data analysis included unsupervised and supervised methods (**Figure 1**). The former were used to establish whether the lipidomics data contained structures that supported prior classifications into baseline versus post-treatment samples, or into subjects with or without neuropathy after treatment. Supervised methods were then used to identify lipid mediators that carried information relevant to this class structure of the data set.

Following log-transformation and missing-value imputation, the lipid marker concentrations were analyzed using unsupervised methods to assess whether they contained structures that were consistent with the prior class structure. Z-standardized data were projected from the high-dimensional space onto lower dimensional planes by means of principal component analysis (PCA) (32, 33). However, among limitations of PCA is that it focuses on the dispersion (variance) of the data, while clustering/subgrouping attempts to identify concentrations (neighborhoods) within the data, making PCA and clustering opposing methods in this sense. Furthermore, PCA fails to separate data sets with non-linear relationships, for which other projection methods have been developed, such as the currently popular t-distributed stochastic neighborhood embedding (t-SNE) (34), which also has limitations and occasionally provides class structure to data sets that do not have class structure (35). Considering these limitations, a second unsupervised approach was used to verify the agreement between the lipidomics data structure and the prior classification, implemented as self-organizing maps (SOM) of artificial neurons (36). In the special form of an “emergent” SOM (ESOM (37)), the present map consisted of 4,000 neurons arranged on a two-dimensional toroidal grid with 50 rows and 80 columns (38, 39). ESOM was used because it has been repeatedly shown to correctly detect subgroup structures in biomedical data sets comparable to the present one (37, 40, 41). The core principle of SOM learning is to adjust the weights of neurons based on their proximity to input data points. In this process, the best matching unit (BMU) is identified as the neuron closest to a given data point. The adaptation of the weights is determined by a learning rate (η) and a neighborhood function (h), both of which gradually decrease during the learning process. Finally, the groups are projected onto separate regions of the map. On top of the trained ESOM, the distance structure in the high-dimensional feature space was visualized in the form of a so-called U-matrix (42, 43), which is the canonical tool for displaying the distance structures of input data on ESOM (38). The visual presentation facilitates data group separation by displaying the distances between BMUs in high-dimensional space in a color-coding that uses a geographical map analogy, where large “heights” represent large distances in feature space, while low “valleys” represent data subsets that are similar. “Mountain ranges” with “snow-covered” heights visually separate the clusters in the data (44). Further details about ESOM can be found in (43).

After verifying that the lipidomics data structure supported the prior class structure into day 1 / 2 or neuropathy/non-neuropathy at day 2, supervised analyses were performed to determine if the lipid mediators were informative for this class structure (feature selection (45)). Again, a mixture of experts approach (29–31) was adopted, including binary logistic regression as a widely used standard (46) and additional machine-learning algorithms known to work well for tabular numerical data were applied, i.e., random forests (47, 48) as a robust tree-based bagging classifier, and support vector machines (SVM) (49) as a hyperplane separation-based method. Classifier tuning is described in the supporting information.

Feature selection was again implemented as a mixture of experts, using 17 different methods, including univariate and multivariate types listed in the Supplementary Appendix. The combination of univariate and multivariate methods is based on the recognition of the limitations of both. Univariate methods, such as standard effect size measures, are commonly used in the analysis of omics data, but it has been shown that they may fail to recognize the high dimensionality of the dataset and inadequately reduce it to a multiple unidimensional problem. As demonstrated elsewhere (50), higher significance in univariate group comparisons does not automatically mean stronger predictive power, and variables with strong predictive power may sometimes be non-significant. Therefore, multivariate feature selection approaches were added; however, acknowledging that the statistical and machine learning models used to infer feature importance also have their limitations. For example, binomial or multinomial regression analysis works only on linearly separable data sets (51), but it is basically impossible to judge whether a given real-world data set is linearly non-separable, support vector machines assume that the data set is separable on hyperplanes, and random forests, assume that the data is separable along the axes of the coordinate system.

After quantifying the importance of each feature by each method, the most relevant subset of lipid mediators was identified by subjecting the importance measure to computed ABC analysis (cABC analysis) (52). This is an item categorization technique that aims to divide a set of positive numerical data into three disjoint subsets labeled “A,” “B,” and “C”. Subset “A” should contain the “few important” elements (53) and its members were therefore retained as candidate features. The number of placements of a mediator in ABC subset A was obtained across all 17 algorithms and subjected to further cABC analysis to find those lipid mediators that were most consistently placed in ABC subset “A” across all 17 feature selection methods. To further reduce the final number of relevant lipid mediators, the cABC analysis could be run recursively as described in (54).

The resulting final sets of lipid mediators were then used to train the algorithms to perform the task of assigning a sample to either the pre- or post-therapy day or to a patient with or without neuropathy in the hold-out 20% validation sample mentioned above. All supervised analyses were performed using cross-validation and training, testing, and validation splits of the dataset as a machine learning standard. That is, a class-proportional random sample of 20% of the dataset was set aside before the analyses started and served as a validation sample that was not touched during algorithm training and feature selection. In the remaining 80% of the dataset, feature selection was performed using supervised analyses in 100-fold nested cross-validation scenarios.

### Calcium Imaging with primary sensory neurons

Primary sensory neurons were cultured as described previously (55). For calcium imaging experiments, neurons were stained with Fura-2-AM (Thermo Fisher) for at least 60 min at 37°C and washed afterwards twice with Ringeŕs solution consisting of 145 mM NaCl, 1.25 mM CaCl_2_ × 2H_2_O, 1 mM MgCl_2_ x 6 H_2_O, 5 mM KCl, 10 mM D-glucose, and 10 mM HEPES adjusted to a pH of 7.3. To investigate the effect of SA1P or LPC 24:0 on different TRP channels, sensory neurons were incubated with the lipids for 1 min at a concentration of 1 or 10 µM, respectively. The gold standard agonists for TRPV1 and TRPA1 were capsaicin (200 nM, 20s) and AITC (allyl isothiocyanate, 75 µM, 30s). Fingolimod was used at a concentration of 1 µM and pre-incubated for 1 h prior to measurement. As a positive control, final stimulation with KCl (50 mM, 1 min) was used to depolarize all neurons. All stimulating compounds were dissolved in Ringer’s solution to their final concentrations.

The calcium imaging data were analyzed using descriptive statistics. All calcium imaging data are presented as the mean ± SEM. Normal distribution was confirmed using the Shapiro-Wilk test. For experiments comparing only two groups, unpaired and heteroscedastic Student’s t-tests were conducted following Welch’s correction. When comparing more than two groups, one-way analysis of variance (ANOVA) was used, and for the comparison of more than three groups, two-way ANOVA was conducted. For all statistical analyses of the calcium imaging data, the software GraphPad Prism 9.5 was used. Statistical significance was set at p value < 0.05.

## Results

All patients received paclitaxel as adjuvant or neoadjuvant therapy, without any other potentially neurotoxic substances. Of the 60 patients from our analysis cohort, two were excluded due to rescheduling of paclitaxel therapy. Blood samples were obtained from 31 patients before and after chemotherapy. Twenty lipid marker variables had > 20% missing values. Following exclusion of these patients and variables and imputation, a data matrix for further analyses was obtained, sized 79 × 255 (79 data set instances, samples, and 255 different lipid mediators Figure 1). An overview of the distribution of the lipid marker data is shown in Figure S1. These included 48 samples drawn on day 1 before therapy and 31 samples drawn on day 2 after 12 cycles of paclitaxel therapy. On day 2, n = 17 of the 31 patients had symptoms of neuropathy (54.9%), which is in line with previous clinical reports on occurrence and severity of paclitaxel-induced neuropathy (56, 57). Most patients reported grade 1 neuropathy, although two patients experienced grade 3 neuropathy. Occurrence and degree of neuropathy were monitored 4.5 years after finishing chemotherapy. The neuropathy lasted for several months or, in many cases, still persisted for 4.5 years after chemotherapy at the last examination (Table S1).

### Results of unsupervised analysis of structure in the lipidomics data supporting prior knowledge

An overview of the distribution of the lipid marker data is shown in Figure S1. PCA yielded d = 28 components with eigenvalues > 1, which together explained 93.93% of the total variance in the lipid mediators (Figure 2). The d = 86 lipid mediators that contributed most to the relevant PCs were identified based on the membership to category “A” in the cABC analysis of the weighted variable contributions to each PC (Figure S2). This was carried over as one of the several feature-importance measures to the supervised analyses reported in the next chapter. On the emergent self-organizing map (ESOM, Figure 2a), a clear separation of two clusters was observed, which provided support that the lipid mediators contained a data structure contingent with the prior classification into pre- and posttherapy samples (Fisher’s exact test: p = 0.01054, odds ratio: 3.57 with 95% confidence interval 1.28 - 10.52; Figure 2b). The separation of samples on the ESOM also corresponded, but to a lesser extent, with the occurrence of neuropathy observed at the time of the 2^nd^ blood sample (p = 0.0328, odds ratio 0.16, 95% confidence interval 0.0137 - 1.022).

**Figure 2:**
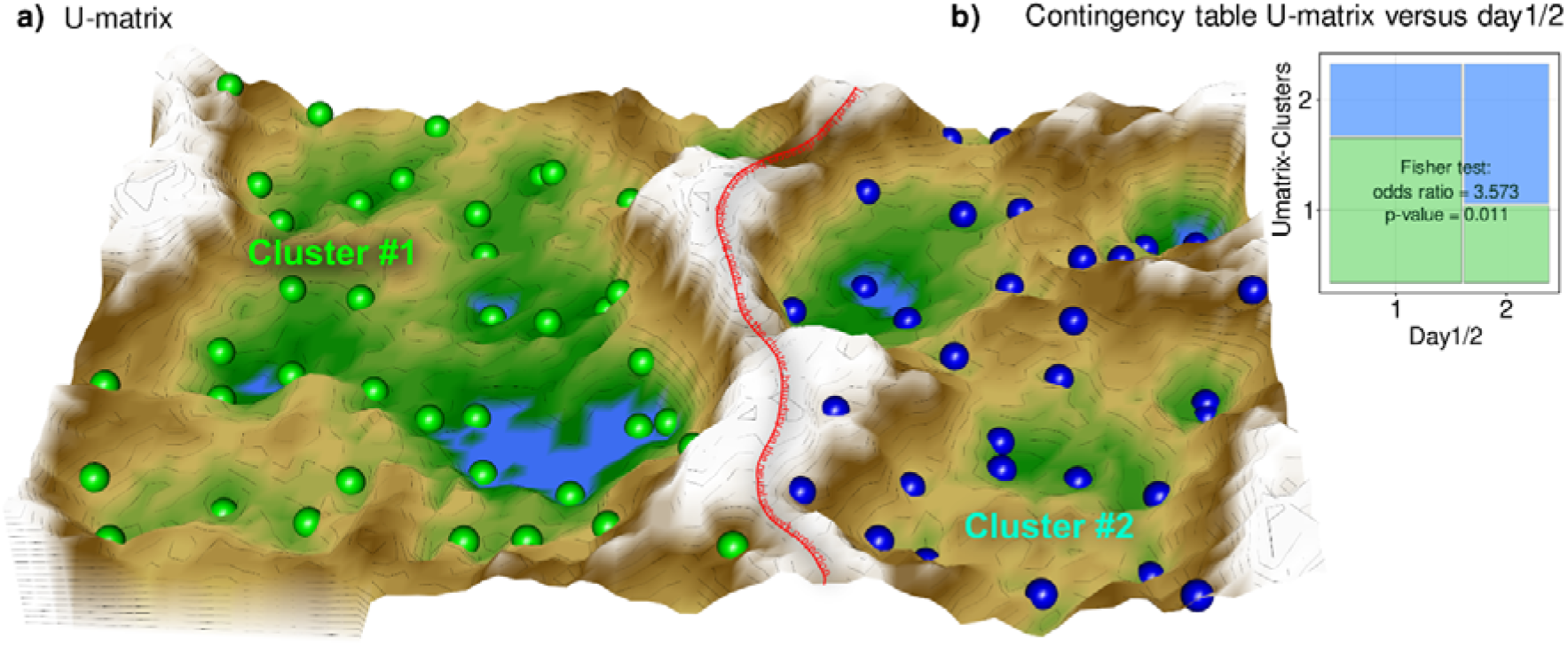
Results of a projection of the z-standardized log-transformed lipidomics data onto a lower-dimensional space by means of a self-organizing map of artificial neurons (bottom). **a):** 3D display of an emergent self-organizing map (ESOM), providing a 3-dimensional U-matrix visualization (78) of distance-based structures of the serum concentrations of d = 255 lipid mediators following projection of the data points onto a toroid grid of 4,000 neurons where opposite edges are connected. The dots represent the so-called “best matching units” (BMU), i.e., neurons on the grid that after ESOM learning carried a data vector that was most similar to a data vector of a sample in the data set. Only those neurons of the originally 4000 neurons are shown that carried vectors of cases from the present data set. Please also note that one BMU can carry vectors of several cases, i.e., the number of BMUs is not necessarily equal to the number of cases. A cluster structure emerges from visualization of the distances between neurons in the high-dimensional space by means of a U-matrix (79). The U-matrix was colored as a geographical map with brown or snow-covered heights and green valleys with blue lakes, symbolizing high or low distances, respectively, between neurons in the high-dimensional space. Thus, valleys (left and right of the “mountain range” in the middle indicate clusters and watersheds, i.e.,, i.e., the line of large distances between neighboring points, indicate borderlines between different clusters. Tat is, the mountain range with “snow-covered” heights separates main clusters according to probe acquisition at day 1 or day 2, i.e., before and after treatment with paclitaxel. BMUs belonging to the two different clusters are colored green or bluish. **b):** Mosaic plot of the prior classes (day 1 or day 2) versus the ESOM/Umatrix based clusters. The separation corresponded to the previous classification into pre- and post-therapy probes (day1/2). Cluster #1 was composed of more probes taken on day #1, while probes from day 2 were overrepresented in cluster #2. The figure has been created using the R software package (version 4.1.2 for Linux; https://CRAN.R-project.org/ (26)), R library “ggplot2” (https://cran.r-project.org/package=ggplot2 (80)) and our R package “Umatrix” (https://cran.r-project.org/package=Umatrix (37)).

The results of the unsupervised analysis thus supported that the lipidomics data contained a structure contingent on a known prior classification. This supported the continuation of data analysis with supervised methods to determine which of the lipids carried relevant information to assign a probe to a particular prior class.

### Results of supervised analyses identifying lipid mediators relevant to the class structure

#### Lipid mediators informative for assigning samples to before or after paclitaxel therapy

Based on the majority vote of the different approaches to feature selection including PCA importance and further univariate and multivariate feature selection methods specified in the supplementary information, d = 77 lipid mediators were found to provide relevant information on whether a sample was collected before or after paclitaxel therapy (Figure S3). When statistical (logistic regression) and machine learning (random forests, support vector machines - SVM) algorithms were trained with this set of lipid mediators, the assignment of a sample to day 1 or 2 was well above the guessing level (Table 2). By contrast, when the training data were randomly permuted, the performance fell to a balanced accuracy of 0.5, i.e., guessing level, which established that the obtained class assignment in the non-permuted scenario had not been due to overfitting. In addition, by rerunning the cABC analysis on the mediators assigned to subset “A” in the first run (“recursive” cABC analysis (54)), the informative set of lipid mediators could be further reduced to d = 27 (Table 1). With these mediators, SVM and random forest were still able to detect whether an instance of a lipidomics dataset was from before or after paclitaxel treatment at a balanced accuracy better than expected from guessing. In summary, with the top hits (“sparse” feature set), three different algorithms (logistic regression, random forests, support vector machines) could be trained to identify, in new cases, whether a blood sample was drawn before or after paclitaxel therapy with a median balanced accuracy of up to 90%.

**Table 1:** Lists of lipid mediators that were most informative in assigning a sample (i) to the first or second sampling time point or (ii) a sample from the second time point to a patient with or without neuropathy. Abbreviations: SA1P: sphinganine-1-phosphate, S1P: sphingosine-1-phosphate, LPE: lysophosphatidylethanolamine, LPC: lysophosphatidylcholine, 2-AG: 2-arachidonoylglycerol, OEA. Oleoylethanolamide.

| **Sample 1 versus sample 2** |  |  |  |  |
| --- | --- | --- | --- | --- |
| **SA1P** | Sphingomyelin 42:1 | Palmitic acid 16:0 | Eicosaeinoic acid 20:1 | PE 38:5 |
| **LacCeramid C16** | LPE 22:6 | Margaritic acid 17:0 | 2-AG | LPC 22:4 |
| **S1P** | LPE 18:0p | Sphingomyelin 42:3 | OEA | Cholesterol sulfate |
| **Sphingomyelin 36:3** | LPE 18:0 | LPC 18:0 | Sphingomyelin 40:1 |  |
| **Ceramide 18:0** | LPC 20:1 | LPC 18:1 | Sphingomyelin 42:2 |  |
| **Ceramide 24:0** | Nervoneic acid 24:1 | Dehydroepiandrosterone sulfate |  |  |
| **Sample 2: neuropathy versus no neuropathy** |  |  |  |  |
| **SA1P** | Sphingomyelin 33:1 | Sphingomyelin 43:1 |  |  |

**Table 2:** Internal validation of the sets of lipid mediators resulting from the feature selection analysis. The different classifiers (linear support vector machine, SVM, random forests, and logistic regression) were trained with subsets of the training data set with all variables (d = 255 lipid mediators as “full” feature set and with the d = 77 or d = 27 lipid mediators that had resulted from the recursive cABC analysis applied on the sum score of selections by 17 different feature selection methods as “reduced” or “sparse” feature sets, respectively. The trained classifiers were applied to a validation sample comprising 20% of the data that had been removed in a class-proportional manner from the dataset at the beginning of feature selection and had not been touched until used in the classifier validation task presented in this table. In addition, the validation task was repeated with training the classifiers with permuted lipid mediators to observe possible overfitting. Shown are the medians and nonparametric 95% confidence intervals (2.5^th^ to 97.5^th^ percentiles) from 5 x 20 nested cross-validation runs. Results of external validation in an independent cohort are shown in the supporting information (Table S3).

| Classifier | Performance measure | Feature set |  |  |  |
| --- | --- | --- | --- | --- | --- |
|  |  | Full | Reduced | Reduced permuted | Sparse |
|  | **Number of lipid mediators** | 255 | 77 | 77 | 27 |
| **SVM** | Balanced accuracy | 0.7 (0.48 - 0.92) | 0.78 (0.61 - 1) | 0.48 (0.23 - 0.76) | 0.75 (0.54 - 0.91) |
| **Random forests** |  | 0.7 (0.56 - 0.83) | 0.75 (0.58 - 0.85) | 0.46 (0.24 - 0.75) | 0.74 (0.55 - 0.88) |
| **Logistic regression** |  | 0.7 (0.52 - 0.85) | 0.77 (0.58 - 0.92) | 0.48 (0.29 - 0.7) | 0.7 (0.49 - 0.89) |
| **SVM** | roc-auc | 0.88 (0.67 - 1) | 0.95 (0.85 - 1) | 0.48 (0.16 - 0.77) | 0.9 (0.81 - 0.99) |
| **Random forests** |  | 0.86 (0.76 - 0.95) | 0.88 (0.81 - 0.98) | 0.48 (0.21 - 0.81) | 0.9 (0.8 - 1) |
| **Logistic regression** |  | 0.81 (0.64 - 1) | 0.88 (0.75 - 1) | 0.46 (0.16 - 0.79) | 0.86 (0.69 - 0.98) |

### Lipid mediators informative for assigning post-paclitaxel therapy samples to neuropathy

The n = 31 samples from day 2 were probably too small to detect whether a sample was from a patient with neuropathy. Although the three algorithms detected neuropathy in new cases, unseen during training, at balanced accuracy of up to 0.75, while only the guess level of 0.5 was achieved when using permuted data for training, the 95% CI of the performance measures was not separated from guess level. Therefore, multivariate feature selection was not considered a valid approach, since it requires that the algorithms from which the feature importance is read can successful perform their task of class assignment (51). Therefore, univariate methods (Cohen’s d, FPR, FWE) were preferred, as well as a direct hypothesis transfer of the top hits from the above-mentioned day1/2 assessments to neuropathy. Classical statistics consisting of direct group comparisons using Kruskal-Wallis tests (58) were performed. The small set of d = 3 lipid mediators that emerged from all three univariate methods as top hits for neuropathy included sphingolipid sphinganine-1-phosphate (SA1P), also known as dihydrosphingosine-1-phosphate (DH-S1P) sphingomyelin 33:1, and sphingomyelin 43:1. Statistical group comparisons verified that the three mediators differed significantly between samples from neuropathy-positive and neuropathy-negative patients (Figure 3).

**Figure 3:**
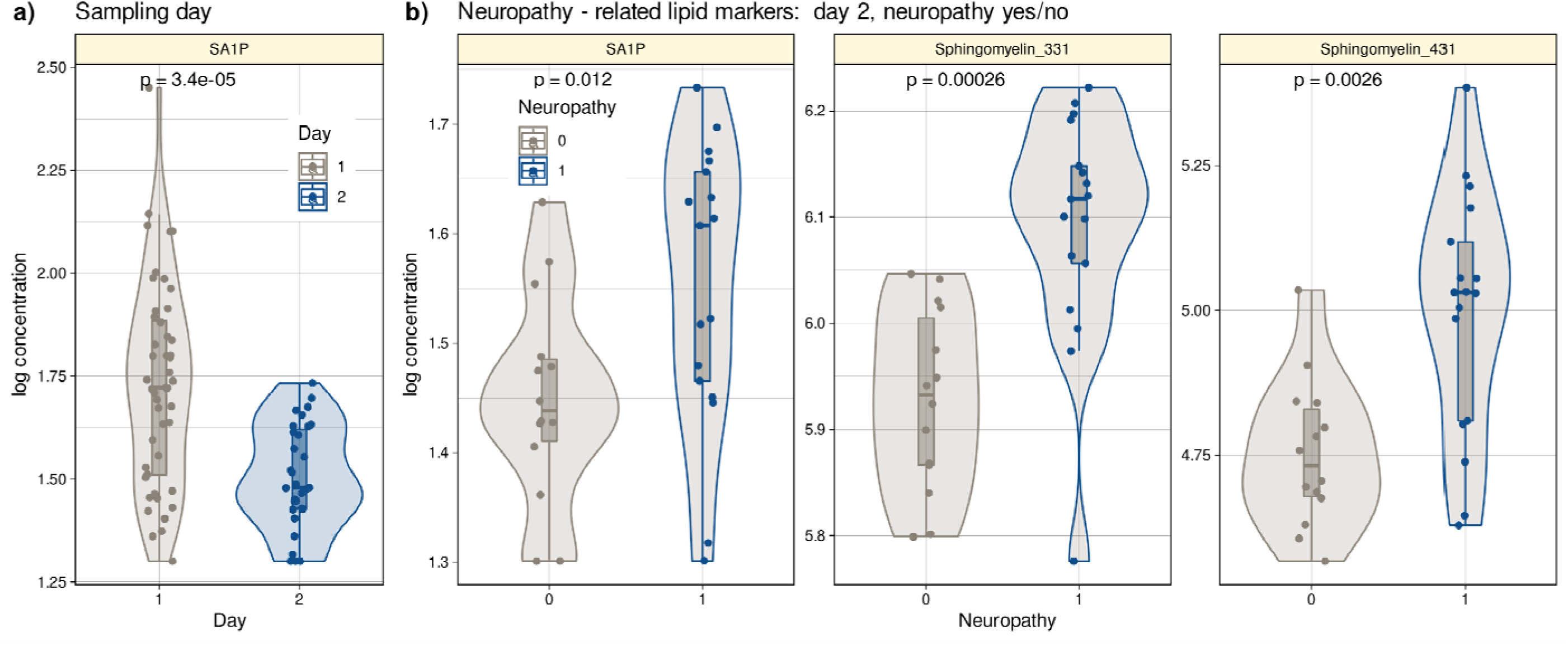
Log_10_-transformed concentrations of lipid mediators shown to be informative for assigning a post-therapy sample to a patient with neuropathy or a patient without neuropathy. Individual data points are presented as dots on violin plots showing the probability density distribution of the variables, overlaid with box plots where the boxes were constructed using the minimum, quartiles, median (solid line inside the box) and maximum of these values. The whiskers add 1.5 times the interquartile range (IQR) to the 75^th^ percentile or subtract 1.5 times the IQR from the 25^th^ percentile. **a):** Concentrations of SA1P (top hit for sample 1 versus sample 2 segregation) are presented separately for the first and second samples. **b):** Concentrations of the top lipid mediators for neuropathy versus no neuropathy in the second sample presented separately for neuropathy-positive and -negative samples. The results of the group comparison statistics (Kruskal-Wallis tests (58)) are given at the top of the graphs. The figure has been created using the R software package (version 4.1.2 for Linux; http://CRAN.R-project.org/ (26)) and the R library “ggplot2” (https://cran.r-project.org/package=ggplot2 (80)).

### Biological in-vitro validation of the machine learning based results

The results of the supervised analysis thus established a limited set of lipid mediators to be regulated in association with paclitaxel therapy or with its side effect of inducing neuropathy. The top hit was sphinganine-1-phosphate (SA1P), providing a basis for in vitro validation of its biological effects in the present context of neuropathy.

Calcium imaging measurements were performed on primary sensory neurons obtained from the murine dorsal root ganglia. We stimulated the neurons with 1 and 10 µM SA1P (ranked as the primary hit) or LPC 24:0 (ranked as one of the least relevant lipids by machine learning analysis). We observed that SA1P caused a direct calcium transient in approximately 11.7% of KCl-responsive sensory neurons (Figure 4a, b). However, LPC 24:0 did not induce any notable activation of sensory neurons at concentration of 1 and 10 µM (Figure S4a-c). To further characterize the SA1P-responding neurons, we investigated their responsiveness to agonists of the TRP channels TRPV1 (capsaicin) and TRPA1 (AITC, allyl isothiocyanate), both of which are hallmarks of subpopulations of primary sensory neurons (59). Stimulating SA1P-responsive neurons with capsaicin and AITC revealed that 73% of these neurons also responded to capsaicin and 25% of them responded to AITC, whereas only 9.6% responded to both stimuli. Neurons were identified as responders to KCl (50 mM, 1 min; Figure 4c, d).

**Figure 4:**
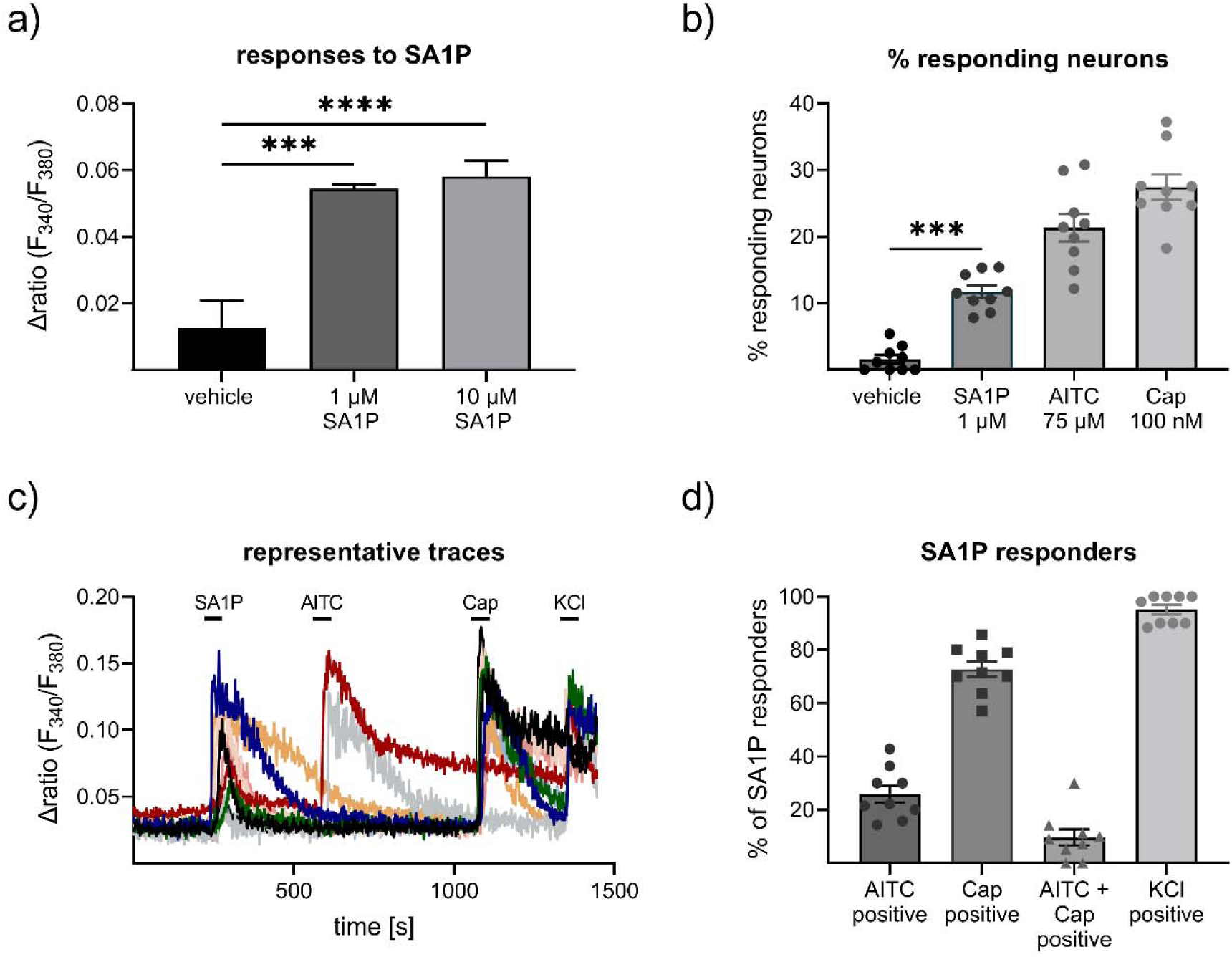
Effects of sphinganine-1-phosphate on primary sensory neurons. a) Neurons were stimulated with SA1P (1 or 10 µM, 1min or vehicle (0.7% methanol (v/v)). b) percentage of responding neurons to vehicle (0.7% methanol (v/v), 1 min), (SA1P (1 µM, 1min), AITC (allyl isothioncyanate, 75 µM, 30s) or capsaicin (caps, 200 nM, 20s). c) representative traces of SA1P-responding neurons and their response to AITC, capsaicin and KCl. d) percentage of SA1P-responding neurons responding to AITC, capsaicin (caps), AITC and capsaicin and KCl. Data are shown as mean ± SEM from at least six measurements per condition with at least 40 neurons per measurement, * p < 0.05, ** p < 0.01, *** p < 0.01, One-way ANOVA.

To identify the receptors or channels responsible for SA1P-mediated calcium transients in sensory neurons, the selective TRPV1 antagonist AMG9810 was used. Neurons were stimulated twice with SA1P (1 µM, 1 min) and AMG9810 (1 µM or vehicle) was added two minutes prior to the second SA1P stimulus. The second SA1P response was entirely abolished when the neurons were treated with AMG9810, but not with the vehicle (Figure 5a-c). The potency of AMG9810 was validated using the same measurement protocol as before but with capsaicin (200 nM, 20s) instead of SA1P, which is the gold standard agonist of TRPV1. AMG9810 completely blocked the second capsaicin response (Figure 5d). The involvement of S1P-receptors previously suggested to be the receptors for SA1P (60), was evident by studying S1PR1 and S1PR3 as the most highly expressed S1P receptors in sensory neurons (61), which are also targets of the approved drug fingolimod. Sensory neurons were incubated with fingolimod or vehicle for one hour and stimulated the neurons with SA1P (Fig. 5e, f). Comparing fingolimod- and vehicle-treated neurons, we observed that the response intensity to SA1P was similar (Figure 5g), while the number of neurons responding to SA1P was significantly decreased after fingolimod treatment (Figure 5h).

**Figure 5:**
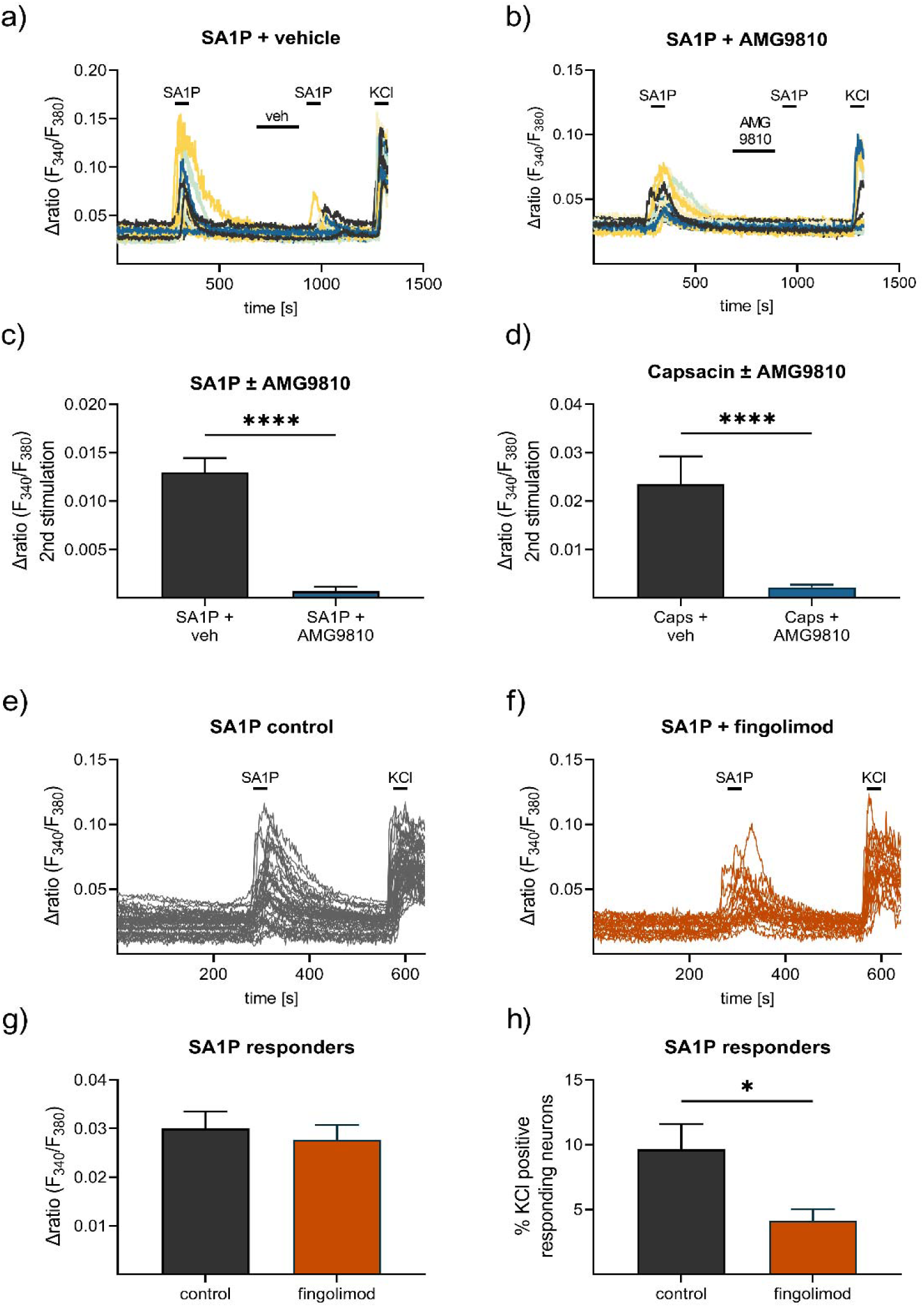
Contribution of TRPV1 and S1P receptors to SA1P-mediated calcium-influx in sensory neurons. Sensory neurons were stimulated with SA1P twice (1 µM, 1 min) and either **a)** vehicle (DMSO 0.003% (v/v), 2 min) or **b)** the TRPV1 antagonist AMG9810 (1 µM, 2 min) prior to the second SA1P stimulus. Cells were depolarized with KCl (50 mM, 1 min) at the end of each experiment. **c)** Statistical analysis of the amplitude of SA1P-mediated calcium transients in sensory neurons treated with either vehicle or AMG9810 (blue). **d)** Statistical analysis of the amplitude of capsaicin-mediated calcium transients (100 nM, 20s) in sensory neurons treated with either vehicle or AMG9810 (blue). **e), f)** Sensory neurons were stimulated with SA1P after preincubation with the S1P1 receptor modulator fingolimod (1 µM, 1 h) or control. **g)** Statistical analysis of the amplitude of SA1P-mediated calcium transients (1 µM, 1min) in sensory neurons treated with either vehicle or fingolimod (1 µM, 1h, orange). **h)** Statistical analysis of the number of SA1P-responding neurons (as % of KCl-positives) after treatment with either vehicle or fingolimod (1 µM, 1h, orange). Data represents mean **±** SEM from at least five measurements per condition with at least 25 neurons per measurement, * p < 0.05, ** p < 0.01, *** p < 0.01, Student’s t-test with Welch’s correction.

### Support of the main results in an independent second patient cohort

The study lacked a separate validation cohort with similar data to the main cohort. However, an independent second cohort was available from another hospital (Oncological Center in Offenbach, Germany). This cohort consisted of 28 patients treated with the “paclitaxel weekly” regimen (paclitaxel 80 mg/m², once weekly) as adjuvant or neoadjuvant therapy for breast or ovarian cancer. All patients provided informed consent into study participation and publication of the results. Blood samples were available from routine collections and were analyzed by LC-MS/MS (Figure S5, Table S3). In contrast to the main cohort, plasma from patients in the second cohort was collected after 6 cycles of paclitaxel treatment due to local routines. Therefore, the second cohort cannot be considered a state-of-the-art validation cohort. In addition, only six patients in this cohort had neuropathy after chemotherapy (26.5%), all with grade 1 neuropathy (Table S4). Despite these limitations, algorithms trained with lipid information from cohort 1 were able to successfully identify whether a probe was taken before or after paclitaxel therapy in the second cohort at a better than guessing level. Furthermore, a trend towards different SA1P concentrations in the plasma of patients after paclitaxel treatment was observed (p = 0.086, Figure 6).

**Figure 6:**
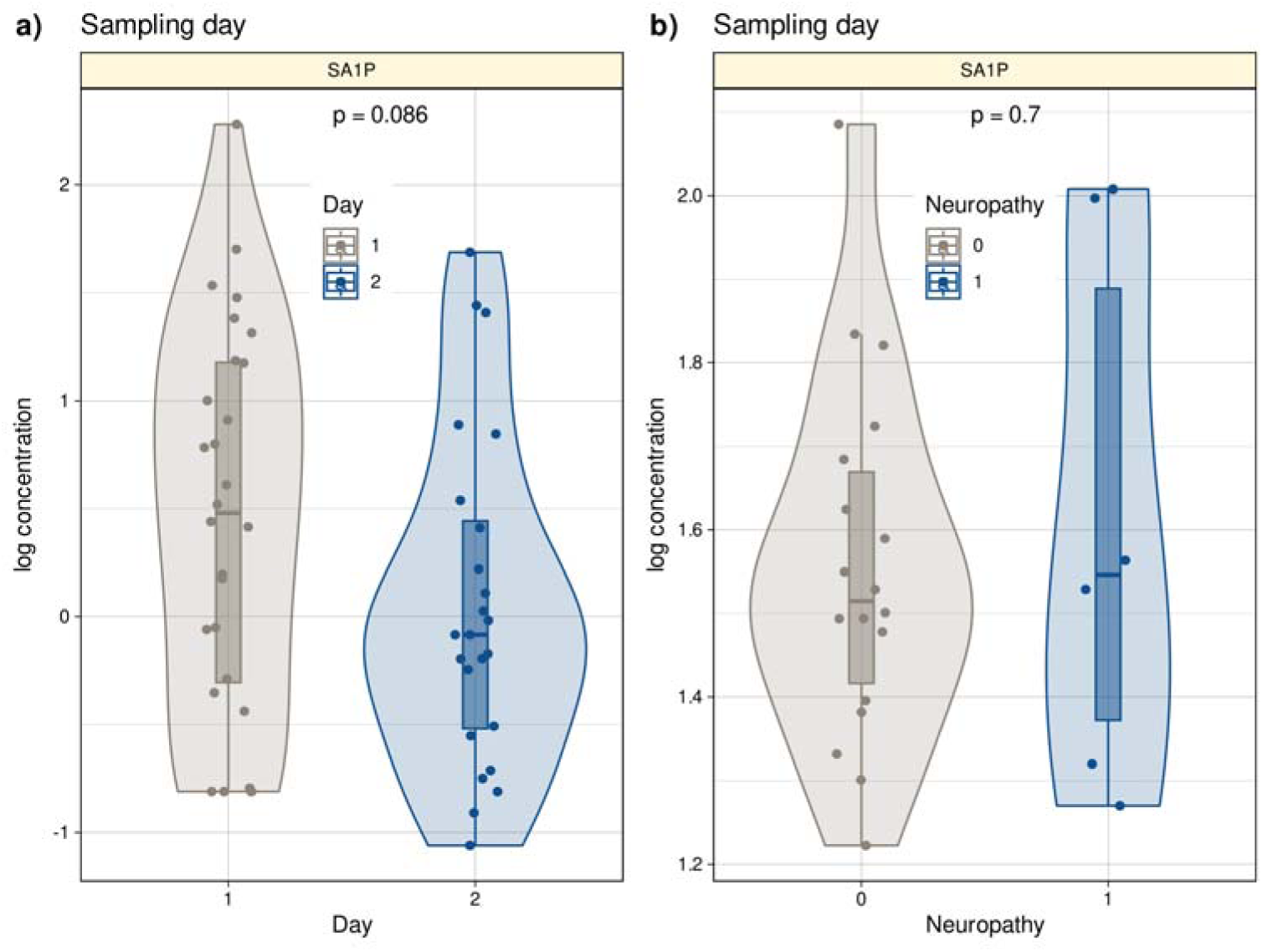
Log10-transformed concentrations of SA1P in the second patient cohort. Individual data points are presented as dots on violin plots showing the probability density distribution of the variables, overlaid with box plots where the boxes were constructed using the minimum, quartiles, median (solid line inside the box) and maximum of these values. The whiskers add 1.5 times the interquartile range (IQR) to the 75th percentile or subtract 1.5 times the IQR from the 25th percentile. a): Concentrations of SA1P (top hit for sample 1 versus sample 2 segregation) are presented separately for the first and second samples. b): Concentrations of S1AP in the second sample are shown separately for neuropathy-positive and -negative samples. Day 1 represents the timepoint before starting chemotherapy. Day 2 represents the timepoint after 12 cycles of paclitaxel chemotherapy. The results of the t-test group comparison statistics are given at the top of the graphs. The figure has been **cre**ated using the R software package (version 4.1.2 for Linux; http://CRAN.R-project.org/ (**26**)) and the R library “ggplot2” (https://cran.r-project.org/package=ggplot2 (**80**)).

## Discussion

For paclitaxel, therapeutic doses range from 80 – 225 mg/m². As CIPN symptoms are dose-dependent, the number of PIPN patients that receive a high paclitaxel dose is higher than the number of PIPN patients receiving a low dose. The cumulative threshold dose above which paclitaxel causes sensory neuropathy is 300 mg/m² (5). In our study, we mainly used a low dose paclitaxel, because this therapeutic regimen is the most widely used paclitaxel monotherapy. Previous studies report an occurrence of neuropathy with this therapeutic regimen is around 50-70%, and most patients (80-90%) are expected to experience Grade 1 neuropathy after 12 weeks (1–3). Our results are within the range reported by these previous studies.

SA1P induced a direct calcium transient in sensory neurons dependent on sphingosine 1-phosphate receptors (S1PR) and the transient receptor potential vanilloid 1 (TRPV1) channel. The results suggest that lipids are altered during paclitaxel treatment and that alterations in sphingolipid metabolism may be critical for the development of paclitaxel-induced peripheral neuropathy in patients. The final (“sparse”) set of lipids regulated between sampling days (before and after paclitaxel treatment) was enriched for sphingolipids. Specifically, sphingolipids were significantly overrepresented among the top hits of lipids regulated between sampling days (Fisher’s exact test: p = 0.01), i.e., while 46 of the original 255 lipid mediators were sphingolipids (18%) (Table S2), 11 of the 27 members of the final sparse marker set (40.7%) belonged to the group of sphingolipids. The main pathway in which the top hits are involved is shown in Figure 7. The classification of patients with regard to neuropathy after paclitaxel treatment was reflected in lipidomics in another sphingolipid, i.e., dihydrosphingosine sphinganine-1-phosphate (SA1P), which was elevated in patients with neuropathy.

**Figure 7:**
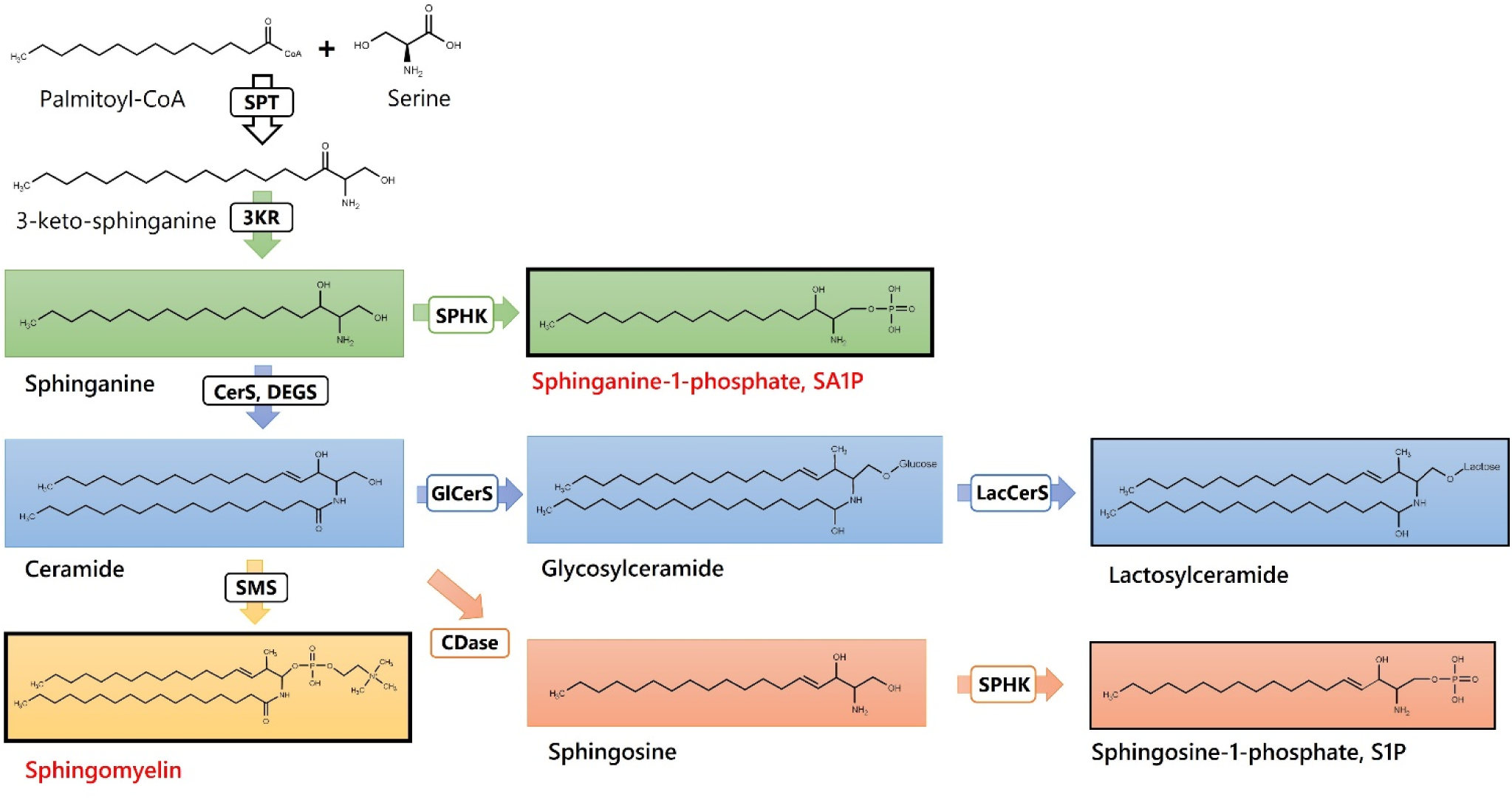
Sphingolipids and Ceramides. (SPT: Serine palmitoyl-transferase; 3KR: 3-ketosphinganine reductase; SPHK: Sphingosine kinase; CerS: Ceramide synthase; DEGS: Dihydroceramide desaturase, GlCerS: Glucosylceramide synthase; LacCerS: Lactosylceramide synthase; SMS: Sphingomyelin synthase; CDase: Ceramidase). Structures were drawn with ChemDraw 20.

The results from an unbiased machine-learning-based analysis are in line with those of previous reports that had used classical statistics mainly. For example, S1P was elevated in the spinal cord of mice after bortezomib treatment and during bortezomib-induced neuropathic pain. Blocking S1P1 receptor S1P1R with fingolimod effectively reduced bortezomib-induced mechanical hypersensitivity in vivo (16). Interestingly, targeting the S1P-S1P1R-axis was also found to reduce paclitaxel-induced neuropathic pain in vivo in a preclinical study (62). Further agreements of the present results relate to previous preclinical reports highlighting the significance of the sphingolipid pathway in persistent and neuropathic pain states (16, 63, 64). In addition, the S1P signaling axis was observed to be relevant in neuropathy and chemotherapy-induced neuropathic pain, which led to the suggestion of targeting S1P receptors as a novel approach to ameliorate chemotherapy-induced neuropathy and neuropathic pain (62, 65–68). Taken together, present results from lipid screening and unbiased machine-learning approach point towards a relevant contribution of sphingolipid signaling in paclitaxel-induced neuropathy in patients, mainly via sphinganine-1-pshophate and sphingomyelins 33:1 and 43:1.

Sphinganine is a key branching point in the sphingolipid pathway, where it can either be acylated to form dihydroceramides or phosphorylated to SA1P by sphingosine kinases (69). Accumulation of sphinganine has been previously associated with reduced activity of ceramide synthase CerS2 (70). Interestingly, low CerS2 expression is a hallmark of various tumors (71). It is conceivable that a subgroup of the patient cohort still exhibits low CerS2 expression after paclitaxel treatment, which is associated with higher SA1P levels and a higher occurrence of neuropathy. We also identified SA1P as a potential proalgesic lipid mediator, as it causes direct calcium transients in approximately 10% of sensory neurons. This effect is mediated, at least in part, by S1P receptors S1P1 and S1P3 and the TRPV1 channel. The TRPV1 channel has previously been identified as an important mediator of neuropathic pain and is associated with exacerbated activity of sensory neurons during paclitaxel-induced neuropathic pain (72–74). Our data indicate that an S1P receptor modulator, such as fingolimod may be a potential treatment strategy for reducing the proalgesic effect of SA1P and possibly for reducing paclitaxel-induced neuropathy in patients.

Several other lipids were found in the extended list of hits previously associated with chemotherapy-induced neuropathy or acute pain in preclinical studies, such as LPC 18:1, sphingosin-1-phosphate (S1P), and 9,10-EpOME. LPC 18:1 has previously been identified as an endogenous activator of TRPV1 and TRM8 and was found at elevated levels in murine DRGs 24 h after oxaliplatin treatment. This may contribute to oxaliplatin-induced acute pain (75). Similarly, 9,10-EpOME was elevated in the DRGs of paclitaxel-treated mice. locking its synthesis with the CYP2J2-inhibitor and the approved drug telmisartan was shown to reduce acute paclitaxel-induced mechanical hypersensitivity in vivo and prevent paclitaxel-induced mechanical allodynia by pretreatment (15). In addition, the direct TRPV1 agonist LPA 18:1 was found in the extended list of hits. Lipid was shown to bind to the C-terminal binding site of TRPV1 to increase the opening probability of the channel (76). Other signaling lipids with potential proalgesic effects and potential TRP channel activators that have been described previously have also been identified as crucial for group separation by our unbiased machine-learning approach, which strengthens the presumption that the identified lipids may indeed be connected with paclitaxel neurotoxicity and paclitaxel-induced neuropathy in patients. Additionally, we found several precursor fatty acids in the extended list of hits, including arachidonic acid, linoleic acid, and palmitic acid. These results imply a major dysregulation of lipids after paclitaxel treatment, leading to enhanced plasma levels of precursors for eicosanoids and oxidized linoleic acid metabolites, which may explain the observed enhanced concentrations of the eicosanoids of 5-HETE, 5,6-DHET, and the oxidized linoleic acid metabolites 9,10-EpOME, 9- and 13-HODE.

Several limitations need to be addressed. First, the size of our patient cohort (n=60 patients, n=31 patients who provided blood samples before and after paclitaxel chemotherapy) allowed for errors due to interindividual differences. Second, the paclitaxel treatment regimen differed in some patients included in this study. While the majority of the patients received paclitaxel as “pacli weekly” regime consisting of a weekly dose of 80 mg/m² for 12 consecutive weeks in, some few patients received paclitaxel doses up to 220 mg/m² or a combination chemotherapy consisting of carboplatin/paclitaxel. These variabilities within the patient cohort hamper interindividual correlations between plasma lipid mediator concentrations and paclitaxel-induced neuropathy throughout the treatment course. Third, the assessment of peripheral neuropathy in patients was performed according to the guidelines of the NCI Common Terminology Criteria for Adverse Events (CTCAE) v5.0, which ranks the severity of neuropathy into five grades but is rather focused on general adverse events of chemotherapy rather than specifically assessing peripheral neuropathy in detail. We did not perform any neurological testing of sensory parameters, such as quantitative sensory testing (QST) (77), to determine the sensory status quo of the patients.

The identification of sphinganine-1-phosphate (SA1P) as a key lipid marker delineating the effects of paclitaxel was based on a comprehensive analysis performed by a “mixture of experts ” of machine learning and classical statistical methods, coupled with in vitro molecular experiments performed on sensory neurons. A rigorous validation process was applied to ensure that the results did not depend on the properties of any single statistical or AI model. This included verifying that the lipidomics data intrinsically supported the existing class structure (days 1/2) using two different data projection methods (PCA, neural network). In addition, the identification of the most relevant lipids associated with past exposure to paclitaxel relied on univariate and multivariate feature selection, and the results were verified by testing three different algorithms that finally could successfully identify from the concentrations of the top hits whether a blood sample was drawn before or after paclitaxel therapy. In addition, all data analyses were subjected to rigorous cross-validation. Results were first validated on a sample split from cohort #1 prior to any feature selection and classifier training, and again on the data from cohort #2. Then, the leading hit from machine learning, SA1P, was validated as a highly active mediator in a neuronal cell model, while a lipid maker deemed irrelevant by machine learning actually showed ineffectiveness in the cell model. The mechanistical details of altered sphingolipid metabolism reflected in their plasma concentrations of paclitaxel patients need to be investigated by further studies.

## Conclusions

Here, we demonstrate that the combination of state-of-the-art lipidomics using LC-MS/MS, LC-QTOF-MS, and machine learning-based data analysis can robustly lead to the generation of testable hypotheses and the identification of biologically relevant signaling mediators of neuropathy in an unbiased manner. Lipidomic profiles were compared within the same patients, allowing analysis of individual paclitaxel-induced lipidome changes in the same patients. These analyses led to the identification of a lipid mediator that can directly activate calcium transients in sensory neurons, thereby modulating nociceptive processing and sensory neuron activity. The identified SA1P, through its receptors, may provide a potential drug target for co-therapy with paclitaxel to reduce one of its major and therapy-limiting side effects.

## Data availability

Data are available on request from the senior author. Relevant Python and R code is available at https://github.com/JornLotsch/PaclitaxelNeuropathyProject.

## Author Contributions

MS and GG conceptualized and designed the experiments. JL and SM performed the machine learning analysis. LH, CA, YS, ST, DT, and NFB performed lipidomics measurements and analyzed the data. SW and MS performed calcium imaging experiments. KG, CB, and CS assembled the patient cohorts. JL, KG, SW and MS wrote the manuscript. All authors have reviewed and edited the manuscript.

## Acknowledgments

MS was supported by the Deutsche Forschungsgemeinschaft (German Research Foundation, DFG, Grants SFB1039 A09 and Z01) and from the Fraunhofer Foundation Project: Neuropathic Pain as well as the Fraunhofer Cluster of Excellence for Immune-Mediated Diseases (CIMD). This work was also supported by the Leistungszentrum Innovative Therapeutics (TheraNova) funded by the Fraunhofer Society and the Hessian Ministry of Science and Arts. JL was supported by the Deutsche Forschungsgemeinschaft (DFG LO 612/16-1). These public funders had no role in study design, data collection and analysis, decision to publish, or preparation of the manuscript. We would also like to thank Drs Tabea Osthues, Béla Zimmer and Vittoria Rimola, as well as Mr. Maksim Sendetski for their help in patient sample processing.

## Supplementary data captions

Figure S1: Log10-transformed concentrations of the analyzed lipid mediators presented as violin plots showing the probability density distribution of the variables; Figure S2: Results of a projection of the z-standardized log-transformed lipidomics data onto a lower-dimensional space by means of PCA; Figure S3: Identification of the lipid mediators that were most informative in assigning a sample to the pre- or post-therapy time point. Feature selection by 13 different methods; Figure S4: Effects of Lysophosphatidylcholine 24:0 (LPC 24:0) on primary sensory neurons; Figure S5: Plasma lipids from the independent second patient cohort. Table S1: Patient characteristics of the 31 patients that gave blood samples before and after chemotherapy from the patient cohort; Table S2: Complete list of lipid mediators included in the analyses, separated by group of lipid and detection method; Table S3: External validation of the classifiers; Table S4: Patient characteristics of the 28 patients from the second cohort that gave blood samples before and after the sixth cycle of chemotherapy. Supplementary methods: data analysis, data preprocessing, supervised and unsupervised machine learning, lipid mediator informative for assigning samples to before or after therapy in an independent second patient cohort, lipidomics analyses of plasma patient samples (Tables 1-10).

